# Presentation, characteristics, treatments and outcomes of mechanically ventilated patients with COVID-19 in Bulgaria

**DOI:** 10.1101/2021.12.17.21267655

**Authors:** Neda Bakalova, Ivats Natsev, Hristo Damov, Irina Yatsenko, Stefanija Jovinska, Rostislav Enev, Mihail Cholakov, Filip Abedinov, Denitza Kalendjieva

**Affiliations:** Department of Anaesthesiology and Intensive Care – Sveta Ekaterina University Hospital, Sofia, Bulgaria; Department of Cardiac Surgery – Sveta Ekaterina University Hospital, Sofia, Bulgaria

## Abstract

**Background:** The first surge of coronavirus disease 2019 (COVID-19) cases in Bulgaria occurred in the fall of 2020. Here we describe the clinical presentation, patient characteristics, treatments and outcomes of mechanically ventilated COVID-19 patients in a newly formed COVID-19 ICU at a tertiary cardiac center in Sofia, Bulgaria.

**Methods:** This is a retrospective observational study of mechanically ventilated COVID-19 patients admitted to Sveta Ekaterina University Hospital in Sofia, Bulgaria, between November 4th, 2020 and January 6th, 2021. Data were collected from electronic and written patient records and charts.

**Results:** We identified 38 critical care patients admitted with respiratory failure and treated with mechanical ventilation at our COVID-19 ICU during this period. The median age was 66 (IQR 57-76, range 27-89) and 74% were male. Most patients, 36 (95%), had at least one comorbidity. The most common comorbidities were hypertension, valvular heart disease, ischemic heart disease and diabetes mellitus. Overall, 27 (71%) patients had a concomitant cardiac disease other than hypertension and 24% were recent cardiac surgical patients. Inotropic support was required in 29 (76%) patients, renal replacement therapy in 12 (32%) patients and prone positioning and ECMO were used in 5 (13%) and 2 (5%) patients respectively. The median duration of mechanical ventilation was 7.5 (IQR 5-14) days overall and 9 (IQR 6-13) days for survivors. At 30-days 28 (74%) of patients had died. Overall, 32 (84%) patients died in hospital and only 6 (16%) patients were discharged home.

**Conclusions:** During the first major surge of COVID-19 cases in Bulgaria, despite the wave arriving later than in other countries, the healthcare system was largely unprepared. In our setting, mortality in mechanically ventilated patients was very high at 84%. Several factors might have contributed to these results, namely the predominance of cardiovascular comorbidities in our patient population, the strained ICU capacity and the lack of medical personnel.

## Introduction

The SARS-CoV-2 virus was identified in Wuhan, China in December 2019 as the causing agent of the COVID-19 disease [1]. The disease itself manifests diversely with some patients experiencing mild, flu-like symptoms while others progress to life-threatening respiratory failure. The virus quickly spread to other parts of the world causing a global pandemic. During the first wave of the disease in the spring of 2020, Bulgaria was largely spared. Indeed, by the 8^th^ of June 2020 – 3 months after the first laboratory confirmed case in Bulgaria – there were only 2727 registered cases or 3% of all tested. This quickly changed in October 2020 with a sharp increase in daily cases to over 1000 per day and a concomitant rise in hospitalizations. By the 31^st^ of December the total confirmed cases were already 201 220 with 7515 fatalities [2].

Initial reports from China, Italy, New York and the UK in the spring of 2020 showed high mortality rates among critically ill patients [3-7]. At that time the SARS CoV-2 virus and the respective COVID-19 disease were largely unknown and the healthcare systems of the affected countries were unprepared for the surge of critically ill patients. Reports that came later from centers which had time to prepare, reported much better outcomes. Unfortunately, the authorities in Bulgaria did not use their time wisely and when the disease finally hit in the fall of 2020, the healthcare system was not ready.

## Methods

We enrolled patients admitted to our COVID-ICU between November 4th, 2020 and January 6th, 2021 and treated with mechanical ventilation. Only laboratory confirmed COVID-19 cases were included. Patients who were intubated in the setting of cardiac arrest secondary to severe hypoxia – **“**crash intubations**”** - were excluded.

Deidentified patient data were collected from hand-written patient charts and electronic patient records. Data on patient age, sex, comorbidities, laboratory findings, treatment modalities, and outcomes was collected. Descriptive statistical analysis of the data was performed. Continuous variables are presented here as mean, standard deviation, median and interquartile range. Data from categorical variables are presented as total number and percentage. No imputation was made for missing data. Given the retrospective nature of the study and the use of deidentified patient data, the need for individual informed consent was waived by the hospital**’**s ethics committee.

## Results

We identified 38 patients admitted to our intensive care unit between Nov 4^th^ and Jan 6^th^ who required invasive mechanical ventilation as part of their treatment. The median age was 66 years (IQR 57-76) with a range of 27-89 years and 28 (74%) patients were male. Baseline characteristics are summarized in Table 1. Twenty (53%) patients were admitted from home while the rest were either transferred from another hospital 6 (15.8%) or from another ward within the same hospital 12 (31.6%). In 9 (24%) patients the COVID-19 infection was acquired while they were inpatients (8 had open heart surgery and 1 had a transcatheter aortic valve implantation (TAVI). The most common symptoms on admission were shortness of breath, temperature, general malaise, and cough. Two patients were admitted with acute thrombosis – one with acute ST-elevation myocardial infarction and the other with thrombosis of the right popliteal artery – and one patient was admitted with an acute aortic dissection and concurrent COVID-19 infection. He underwent emergency cardiac surgery and was then transferred to the COVID ICU.

**Table 1.** Clinical Characteristics of the Patients on Admission.

| Characteristic | Patients (n = 38) |
| --- | --- |
| Age, mean ± SD (range) | 65 ± 14 (27-89) |
| Sex, no. (%) |  |
| - Male | 28 (74%) |
| - Female | 10 (26%) |
| Body Mass Index, median (IQR)<sup>a</sup> | 27 (25-31) |
| Admission site, no. (%) |  |
| - Home | 20 (53%) |
| - Hospital transfer | 6 (16%) |
| - From another ward within the hospital | 12 (32%) |
| Transmission route, no. (%) |  |
| - Community-acquired | 29 (76%) |
| - Hospital-acquired | 9 (24%) |
| Comorbidities, no. (%) |  |
| - None | 2 (5%) |
| - Hypertension | 24 (63%) |
| - Diabetes mellitus | 12 (32%) |
| - Chronic obstructive pulmonary disease | 3 (8%) |
| - Chronic kidney disease | 11 (29%) |
| - Ischemic heart disease<sup>b</sup> | 14 (37%) |
| - Valvular heart disease<sup>c</sup> | 20 (53%) |
| - Congenital heart disease | 2 (5%) |
| - Cancer<sup>d</sup> | 4 (11%) |
| Total patients after cardiac surgery | 13 (34%) |
| Patients after recent cardiac surgery<sup>e</sup> | 9 (24%) |
| - EuroSCORE II, median (IQR) | 5.72 (4.13-7.50) |
| **Echocardiography findings on admission, n/total n (%)** |  |
| - Normal TTE | 4/34 (12%) |
| - EF < 50% | 13/34 (38%) |
| - Moderate or severe valvular disease | 20/34 (59%) |
| - Raised systolic pulmonary artery pressure $\geq 35$ mmHg | 17/34 (50%) |
| - Pleural effusion | 5/34 (15%) |
| **Admission symptoms and timing for patients with community-acquired COVID (n=29), no. (%)** |  |
| - Cough | 14 (48%) |
| - Fever $> 38$ | 22 (76%) |
| - Shortness of breath | 26 (90%) |
| - Malaise | 22 (76%) |
| - Thrombosis | 2 (7%) |
| - Nausea and vomiting | 1 (3%) |
| - Tachyarrhythmia | 3 (10%) |
| Number of days from symptom onset to admission, median (IQR) | 6 (3-7) |
| Number of days from admission to ICU, median (IQR) | 0 (0-1.5) |
| Number of days from admission to intubation, median (IQR) | 2 (1-3.5) |

<sup>a</sup> Data available for 25 patients
<sup>b</sup> Defined here as previous PCI with stenting or coronary artery bypass
<sup>c</sup> As assessed by TTE on admission – only moderate and severe lesions included
<sup>d</sup> These included: lung cancer with pulmonary resection; breast cancer with lung metastasis; prostate cancer – treated and in clinical remission; colon cancer post-surgery within the same year.
<sup>e</sup> In this admission

Comorbidities were common in this patient group. The most common comorbidity was hypertension (24 patients, 63%), followed by valvular heart disease (20 patients, 53%), ischemic heart disease (14 patients, 37%), and diabetes mellitus (12 patients, 32%). As most patients were either current or previous patients of our hospital, which is a tertiary center for cardiovascular disease, concomitant cardiovascular pathology was present in 27 out of 38 patients. This included ischemic heart disease (defined here as previous CABG or PCI), valvular heart disease (moderate or severe), rhythm abnormalities requiring a permanent pacemaker or ICD device, and congenital heart disease. A substantial number of patients – 9 (24%) – were admitted to the COVID ICU following open heart surgery for elective (4 patients), urgent (4 patients) and emergency procedures (1 patient). Their median EuroScore II was 5.72 (IQR 4.13-7.50). Only two patients (5%) did not report any comorbidities.

Transthoracic echocardiogram (TTE) was performed in 34 (89%) patients on admission. Of those, 13 (38%) patients showed some left ventricular dysfunction defined as EF<50%, 20 (59%) patients had moderate or severe heart valve disease and 17 (50%) patients had raised PA pressure above 35mmHg. Pleural effusions were found in 5 (15%) patients. Only 4 (12%) of the 34 examined patients had a normal TTE on admission.

Laboratory findings are summarized in Table 2. Lymphocytopenia was common with 95% of all patients presenting with a lymphocyte count below 1.5×10^9 per liter. Most patients had their CRP, IL-6 and Procalcitonin levels tested on admission and all were elevated, however, the CRP and IL-6 more so than the Procalcitonin which is predominantly associated with bacterial infections. Almost half of patients had a raised high sensitivity troponin at admission which is in line with previous reports and in accord with the transthoracic echocardiography findings on admission which show impaired LV function in over a third of our patients. The lactate levels on admission were raised above 2.2mmol/L in 54% of patients and the median SOFA score on ICU admission was 5.

**Table 2.** Laboratory, clinical and imaging data on ICU admission and during the course of disease.

| Laboratory data |  | Reference range |
| --- | --- | --- |
| White-cell count |  |  |
| - Median (IQR) - per L | 9.89 (6.50-13.73) | 3.5 - 10.5 |
| - Distribution – no. (%) |  |  |
| ≥ 10.5 /L | 16 (42%) |  |
| ≤ 3.5 /L | 4 (10%) |  |
| Lymphocyte count |  |  |
| - Median (IQR) - per L | 0.625 (0.430 - 0.913) | 1.5 - 2.8 |
| - Distribution – no. (%) |  |  |
| ≤ 1.5 | 36 (95%) |  |
| Lowest platelet count first 3 days in ICU, median (IQR) - x10<sup>9</sup> per L | 128 (95-216) | 150 - 450 |
| CRP on admission, median (IQR) – mg/L | 172 (116-241) | 0 - 5 |
| Procalcitonin, median (IQR) - ng/mL | 1.72 (0.83 - 4.38) | 0.05 - 0.5 |
| IL-6, median (IQR) - pg/mL | 106 (33 – 247) | 0 – 7.5 |
| Aspartate aminotransferase, median (IQR) - U/L | 56 (35-75) | 0 – 40 |
| Alanine aminotransferase, median (IQR) - U/L | 32 (22-48) | 0-40 |
| Total bilirubin, median (IQR) - μmol/L | 9.95 (6.9 - 13.3) | 0 – 21 |
| Creatinine, median (IQR) - μmol/L | 107 (87 – 150) | 74 - 127 |
| Ferritin, median (IQR) - μg/L | 665 (570 – 695) | 20 - 250 |
| Lactate dehydrogenase, median (IQR) - U/L | 536 (353 – 615) | 0 - 248 |
| Fibrinogen, median (IQR) - g/L | 4.1 (3.45 - 5.4) | 2 – 4.1 |
| D-dimer ≥ 0.5 μg/mL - no. (%) | 17 (49%) |  |
| HS Troponin, median (IQR) - ng/mL | 0.0573 (0.017 - 0.175) | 0.0 - 0.0175 |
| HS Troponin levels > 0.03 - no. (%) | 22 (65%) |  |
| Brain-type natriuretic peptide, median (IQR) - pg/mL | 238 (142 – 398) | 0 – 100 |
| Lactate, median (IQR) - mmol/L | 2.6 (1.45 - 3.52) | 0.5 - 2.2 |
| Lactate > 2.2 mmol/L - no. (%) | 20 (54%) |  |
| **Clinical measures** |  |  |
| SOFA score on admission to ICU, median (IQR) | 5 (3 – 8) |  |
| SOFA score at 96 hours, median (IQR) | 7 (4.5-10) |  |
| Acute respiratory distress syndrome |  |  |
| - mild – no. (%) | 8 (21%) |  |
| - moderate – no. (%) | 21 (55%) |  |
| - severe – no. (%) | 9 (24%) |  |
| **Chest radiography findings on admission – no./total no. (%)** |  |  |
| - Bilateral infiltrates | 28/36 (78%) |  |
| - Congestion | 4/36 (11%) |  |
| - Pleural effusion | 3/36 (8%) |  |
| - Clear | 2/36 (6%) |  |
| **Chest CT imaging findings on admission – no./total no. (%)** |  |  |
| - Bilateral ground glass opacifications | 10/12 (83%) |  |
| - Unilateral consolidation | 2/12 (17%) |  |
| - Pleural effusion | 5/12 (42%) |  |

CRP – C-reactive protein; IL-6 – interleukin-6; SOFA score– sequential organ failure assessment score;

A chest radiographic image was obtained in 36 patients on day one and 2 patients had a computer tomography scan instead. In 10 patients both a radiograph and a computer tomography (CT) scan were done. On admission, bilateral infiltrates were seen on (78%) of chest radiographs, 4 (11%) showed congestion, 3 (8%) had pleural effusion and in only 2 (6%) patients the initial chest x-ray was clear. The computer tomography imaging showed bilateral ground glass opacifications in 10 out of 12 patients and unilateral infiltrates in the remaining 2 patients. Pleural effusions were seen in 5 (42%) of the computer tomographic studies.

The ICU management, complications and outcomes are summarized in Table 3. All patients in this case series received mechanical ventilation during their stay. The median duration of mechanical ventilation was 7.5 days (IQR 5-14) and in survivors it was 9 days (IQR 6-13). Acute respiratory distress syndrome (ARDS) was observed in all patients in our case series with 8 (21%) meeting the criteria for mild ARDS, 21 (55%) for moderate and 9 patients (24%) developed severe ARDS by 72 hours after intubation [8]. The worst partial pressure of arterial oxygen to inspired oxygen (PaO2/FiO2) ratios on days 1, 2 and 3 are summarized in Table 3. The median positive end-expiratory pressure was 12 (IQR 10-14) at its highest setting. Five (13%) patients underwent prone positioning. Six (16%) patients had a percutaneous tracheostomy procedure. In 11 (29%) patients non-invasive mechanical ventilation was attempted before intubation. No patients received high-flow nasal cannula support as that modality was not available at our hospital at the time. Two (5%) patients received veno-venous ECMO.

**Table 3.**
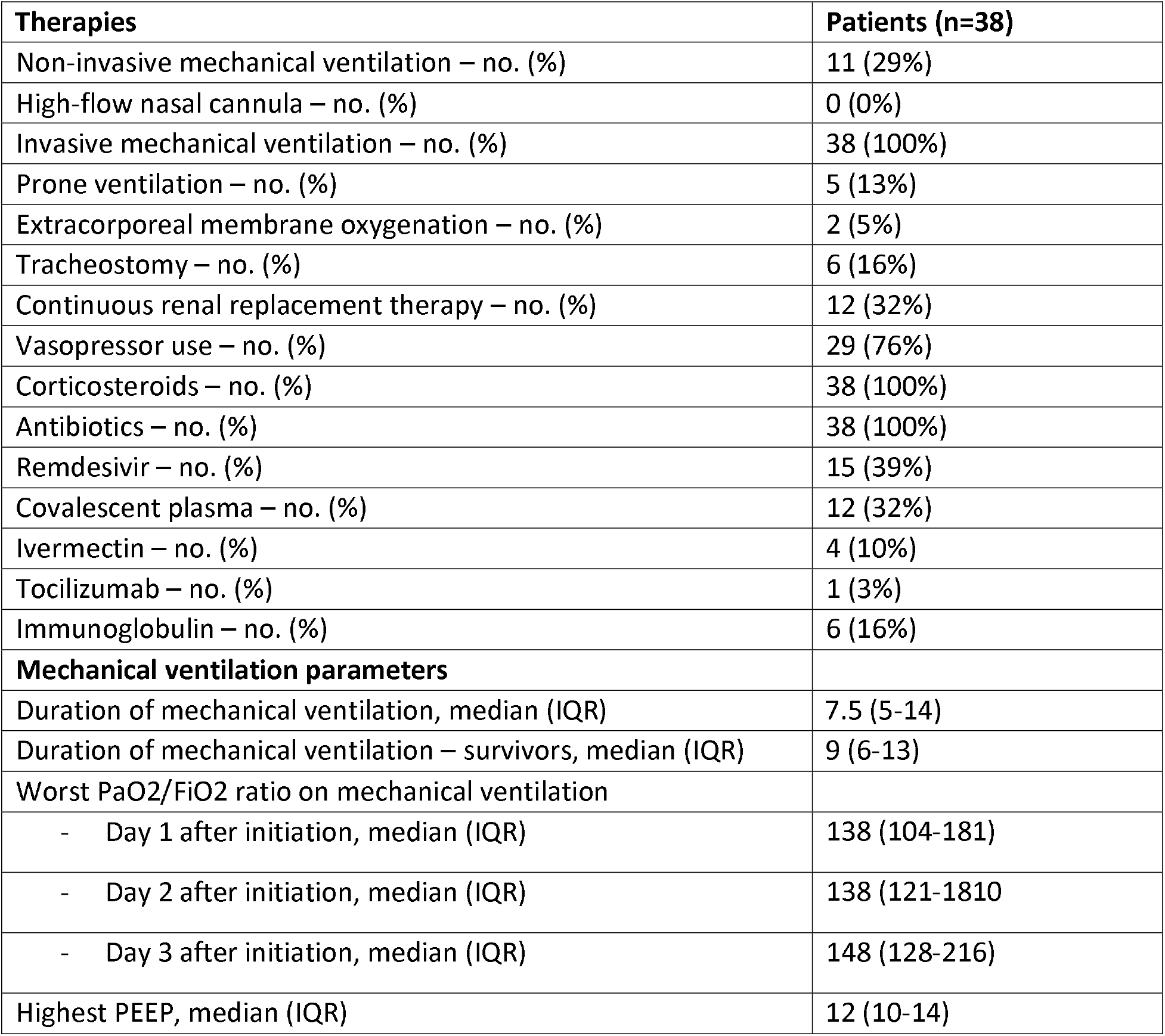
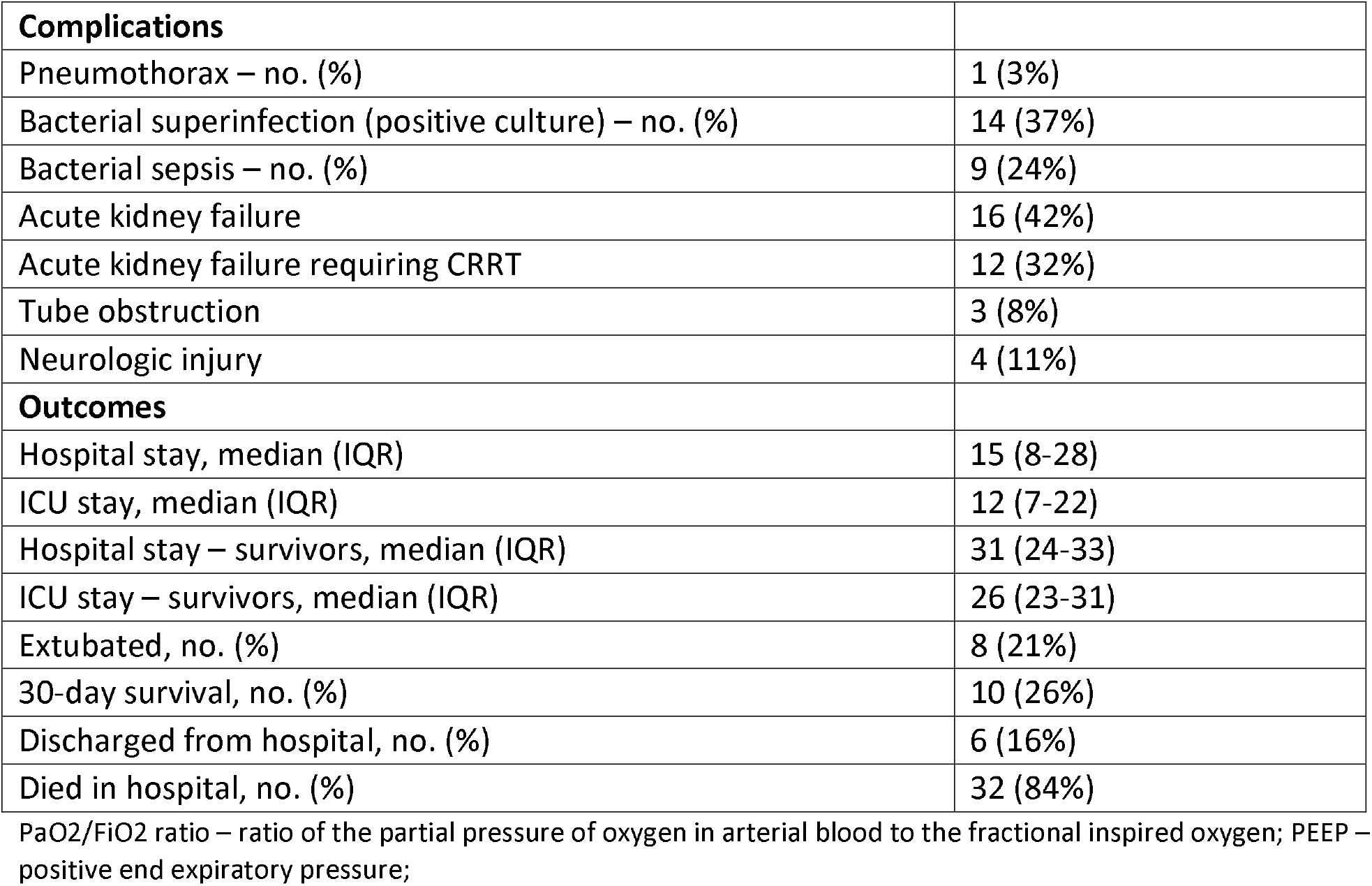
ICU management and clinical outcomes.

Seventy-six percent of patients required vasopressor support during their time in the ICU and roughly a third of all patients required continuous renal-replacement therapy. All patients were given corticosteroids and all received antibiotics. Remdesivir was used in 15 (39%) patients and convalescent plasma was transfused to 12 (32%) patients.

The most common complication in our case series was acute kidney injury with 16 (42%) of patients meeting the RIFLE criteria for kidney failure – increase in serum creatinine more than 3-times the baseline level and/or creatinine above 354 µmol/L with an acute rise of more than 44 µmol/L [9]. In 12 (32%) patients continuous renal-replacement therapy was initiated. The second most common complication was bacterial superinfection with 14 (37%) of patients having a positive blood or respiratory culture taken after at least 72 hours of ICU admission. Nine (24%) patients eventually died with the clinical picture of bacterial sepsis.

30-day mortality was 74% and overall mortality was 84% with only 6 (16%) patients surviving to discharge from hospital. The median age of survivors was 54 years (IQR 49-57; range 45-67) which was 12 years younger than the overall for the case series and 3 (50%) of survivors were women. Only 2 (33%) survivors required vasopressor support and none required renal-replacement therapy. Of the 5 patients who required proning, 4 (80%) died. All 12 (32%) patients who required renal replacement therapy and both ECMO patients died.

## Discussion

This single center case series is to our knowledge the first published report on the clinical characteristics, treatments and outcomes of critically ill COVID-19 patients from Bulgaria. Here we present the data for 38 mechanically ventilated patients admitted to our newly-formed COVID-19 ICU at University Hospital Sveta Ekaterina in Sofia, Bulgaria in the course of 2 months.

Our patients had demographic characteristics similar to previously reported case series from the UK, New York and Italy [5-7]. In terms of accompanying comorbidities (95%) it seems our patient population was sicker compared to data from Wuhan (40%-48%) and Seattle (33%), but comparable to reports from Washington (86%), Lombardy (68%) and Vancouver (73.5%) [3-5, 10-12]. The presence of cardiac comorbidities in our case series, however, is unmatched by previous reports with 71% of our patients having a cardiovascular disease other than hypertension. In comparison, the percentage of cardiac comorbidities from New York, Lombardy, Tokyo, Washington and Atlanta were 11.1%, 21%, 14.2%, 42.9%, and 18.9% respectively [5,6,11,13,14].

We identified the presence of pulmonary arterial hypertension above 35mmHg in 45% (17/38 patients) as measured by trans-thoracic echocardiography on admission. This is a much higher percentage than previously reported [15]. In 5 patients there was a cardiac pathology that could explain the raised pressures. In the remainder of cases the increased pulmonary pressures were likely a result of the acute hypoxemia associated with the COVID-19 infection.

The worst median Pa/Fi ratio within 24 hours of intubation in our patient group was 138 which is comparable to previously reported from Atlanta (Pa/Fi 132), Vancouver (Pa/Fi 180), and Lombardy (Pa/Fi 160) [5,12,14].

We also report on outcomes of patients with severe COVID-19 in the early postoperative period after cardiac surgery. Those were a particularly vulnerable patient group. Many had high EuroSCORE II and their condition was poor even without a concomitant COVID-19 infection and subsequent ARDS. In fact, only 2 surgical patients survived and both had low EuroSCORE II (mean 1.22%), while among the 7 fatalities the mean EuroSCORE II was 9.17%.

The outcomes from this case series are discouraging with an 84% mortality of mechanically ventilated patients. This figure is comparable to the earliest reports from Wuhan, New York, and the UK when reported mortality among ventilated patients with ICU outcome was 86-97%, 76-97% and 67% respectively [3,4,6,7]. It is, however, much higher than data from other centers such as Vancouver, Atlanta, Singapore, Tokyo where mortality among intubated patients was 20.3%, 35.7%, 15.4% and 35.7% respectively [12-14, 16]. Even though all cited reports were observational studies and causality cannot be firmly established, a common denominator in the latter group is the absence of strained ICU capacity and the preservation of the normal provider to patient ratios to pre-pandemic times. In fact, we know that having to repurpose other units for critical care patients and reductions in the provider to patient ratio leads to worse outcomes even in the absence of a pandemic [17-18].

Several reasons may explain the high mortality in our patient group. First, our patients had a high frequency of comorbidities, especially cardiovascular disease. Many were recent cardiac surgery patients. Often, we were not treating single organ failure but both respiratory and cardiac failure. Then there were a number of organizational and staffing challenges that seem to be key to the survival of ICU patients as evident from previous reports. A major difficulty was the structure of the newly formed ICU as it was a repurposed surgical ward and the ICU organization that we were used to could not be followed. Beds were in separate rooms with no central monitoring. Monitoring, in general, was scarce and not all patients were provided with the standard ECG (electrocardiogram), pulse oximetry, and blood pressure monitoring.

Critical care nurse shortages were evident even before the pandemic and became blatantly apparent when a new 16-bed ICU was opened. The nurse-to-patient ratio was 1:5 and even 1:8 with half the nurses being with a non-critical care background. There was no time for them to be trained appropriately for their new role and asking them to take on jobs outside their usual scope of practice led to many avoidable incidents and caused significant mental strain on staff. Personal protective equipment, especially masks, were in short or at best sporadic supply. No fit-testing was done. Not surprisingly infection rates among the medical personnel were close to 100% which meant that all staff at one time or another were on a sick leave instead of taking care of patients.

On a government level, public measures to reduce the spread of the infection were belated and inadequate. In comparison to March-May when disease incidence was low but measures were very tight with a complete lockdown, in October-December no lockdown was established. Planned surgical operations were suspended rather late - on the 25th of October - with many centers continuing to carry out non-urgent surgeries well into the peak of the pandemic and thus additional ICU beds could not be freed in time for the rapidly increasing numbers of critical COVID-19 patients.

### Limitations

Our study has several important limitations. First, it is a single center observational study and all assumptions for a causal link between strained capacity, inadequate staffing, and poor outcomes are strictly theoretical and cannot be firmly established. Our study sample was small in size and our patient population was with an unusually high rate of cardiovascular comorbidities which might have led to a worse survival.

## Conclusions

In our setting, mortality in critically ill patients requiring mechanical ventilation was high and similar to previously published data from the early spread of COVID-19 in countries whose ICU capacity was strained. Our data suggest that patients with concomitant cardiovascular disease are a particularly vulnerable patient population.

## Data Availability

All data produced in the present work are contained in the manuscript

## Acknowledgments

The authors would like to acknowledge all fellow physicians, nurses, medical students and healthcare workers who continued to carry on their duties despite the long shifts, the extra work, the equipment shortages, the lack of personal protective equipment, and the despairing circumstances.

## Author Contributions

Data curation: Neda Bakalova

Formal analysis: Neda Bakalova

Investigation: Neda Bakalova, Ivats Natsev, Rostislav Enev, Hristo Damov, Irina Yatsenko, Stefania Jovinska, Mikhail Cholakov

Methodology: Neda Bakalova

Supervision: Filip Abedinov

Validation: Denitza Kalendjieva

Writing – original draft: Neda Bakalova

Writing – review & editing: Ivats Natsev

